# Antihypertensive medication discontinuation after new-onset stroke

**DOI:** 10.1101/2025.10.06.25337461

**Authors:** Jianian Hua, Guo-Chong Chen, Qingmei Chen

## Abstract

**Backgrounds:** To investigate the proportion of stroke survivors with pre-stroke hypertension who successfully discontinued antihypertensive medication after stroke.

**Methods:** This study analyzed data from 9,513 participants from the English Longitudinal Study of Ageing (ELSA; waves 4, 6, and 8) and the China Health and Retirement Longitudinal Study (CHARLS; waves 1, 2, and 3), all of whom received BP measurements across three waves. The BP control status among stroke survivors were reported according to occurrence of hypertension before and after stroke.

**Results:** Among these, 262 participants experienced a first-ever stroke during follow-up. After stroke, 12.1% to 13.6% of stroke survivors with pre-stroke hypertension maintained controlled BP in the absence of antihypertensive medication. In the ELSA cohort, the proportion of controlled BP after stroke among hypertensive participants was significantly lower in those with new-onset hypertension after stroke (27.3%) compared to those with pre-stroke hypertension (63.6%, *p*=0.023).

**Conclusion:** Under rigorous BP monitoring, a subset of stroke survivors with pre-stroke hypertension may safely discontinue antihypertensive therapy. Furthermore, stroke survivors with hypertension occurring after stroke may be a key target group for improving post-stroke BP control.

## Introduction

Long-term antihypertensive medication is recognized as a cornerstone of secondary prevention for hypertensive stroke survivors to improve prognosis.^1^ However, a number of hypertensive stroke survivors observed that they could maintain optimal blood pressure (BP) in the absence of medication following new-onset stroke. Typically, stroke survivors, particularly those with severe arterial stenosis, are firstly told that their BP requires careful consideration. In such cases, excessively low BP, especially below 90/60 mmHg, may compromise cerebral perfusion and potentially worsen neurological outcomes.^2,3^ Some patients experience an initial post-stroke hypotensive period that leads to temporary cessation of anti-hypertensive medication, and subsequently discover that the medication may not be required. These individuals consider themselves “fortunate” due to the reduced medication burden, although this phenomenon may be attributed to underlying comorbidities, such as atrial fibrillation or heart failure. Nevertheless, to our best knowledge, the true prevalence of stroke survivors who can safely discontinue antihypertensive medication remains unknown. This knowledge gap stems from the limitations of conventional stroke cohorts, which rely on post-stroke BP measurements and self-reported pre-stroke hypertension history, lacking BP data before stroke onset.

The primary objective of this study was to determine the proportion of stroke survivors who could successfully discontinue antihypertensive medication, while the secondary objective was to report BP control status among stroke survivors.

## Methods

The current study employed a retrospective design using data from two nationally representative longitudinal cohorts: the English Longitudinal Study of Ageing (ELSA) and the China Health and Retirement Longitudinal Study (CHARLS).^4,5^ We used data from wave 4 (2008), wave 6 (2012), and wave 8 (2016) of ELSA, along with wave 1 (2011), wave 2 (2013), and wave 3 (2015) of CHARLS, as BP measurement was available in these waves. Ethical approval for ELSA was approved by the London Multicentre Research Ethics Committee (MREC/01/2/91), and CHARLS obtained approval from the Institutional Review Board of Peking University (IRB00001052-11015 for household survey). We finally included 2,396 participants from ELSA and 7,117 participants from CHARLS who had complete BP data in all three waves and no stroke history at baseline (Figure 1 and Table S1).

**Figure 1.**
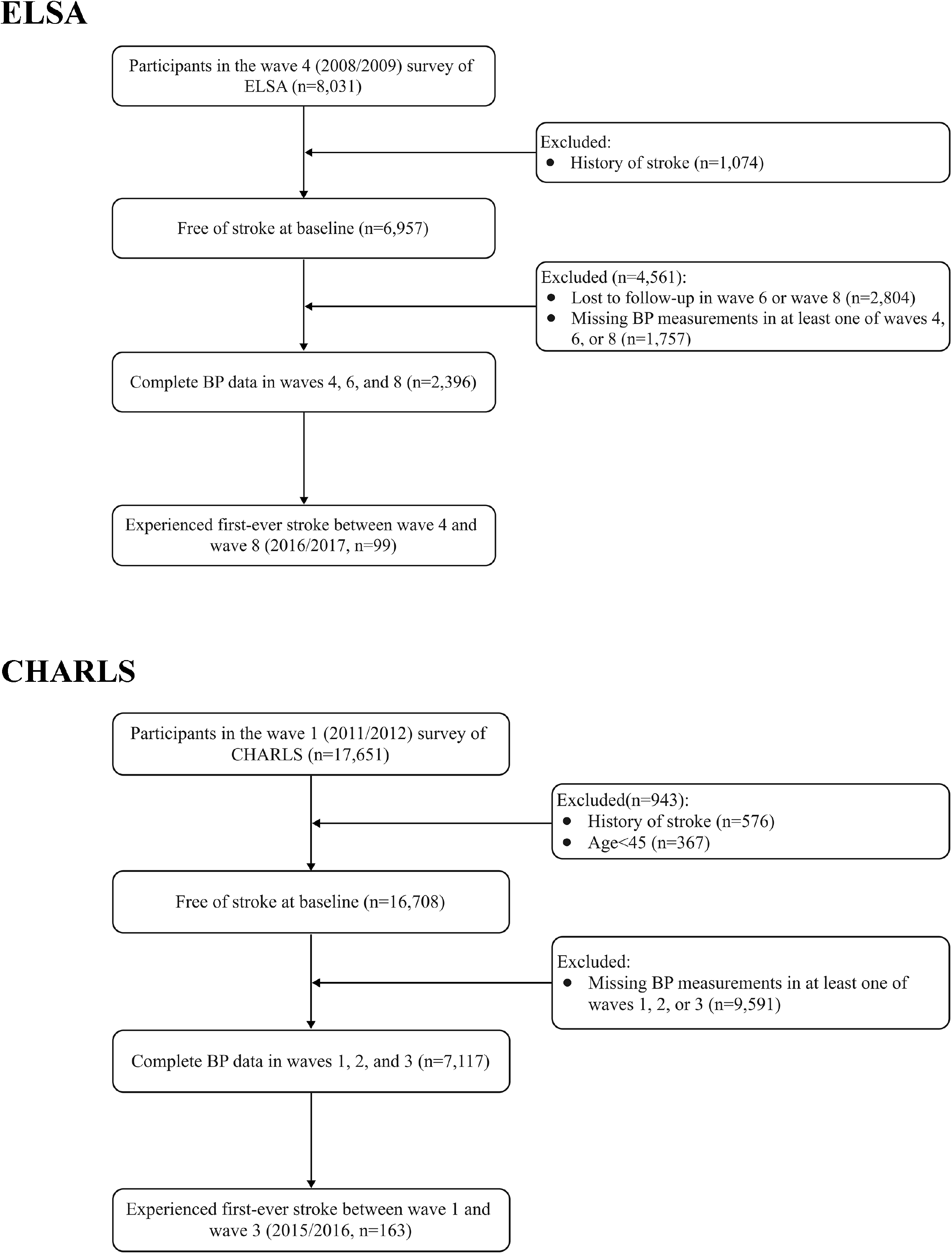
Derivation of the study sample.

Stroke was identified by self-reported physician diagnosis.^6^ Prior analysis reported that self-reported stroke diagnosis does not differ greatly from physician-verified strokes.^7^ The average BP from two or three BP readings was used for analysis. Hypertension was defined as BP measurement ≥ 140/90 mmHg or use of antihypertensive medication.^8^ Controlled BP was defined as systolic BP<140 mmHg and diastolic BP<90 mmHg.

Descriptive statistics were used to characterize the proportion of these individuals. All analyses were conducted using SAS 9.4 (SAS Institute, Cary, NC, USA). A two-sided *p* value of less than 0.05 was regarded as statistically significant.

## Results

During the follow-up of 8 years for ELSA and 4 years for CHARLS, 262 participants experienced their first-ever stroke and had BP measurements across all three surveys, including 99 participants from the ELSA cohort (mean [SD] age, 69.3 [7.0] years; 55.6% male) and 163 participants from the CHARLS cohort (mean [SD] age, 61.0 [8.5] years; 48.5% male, Table 1).

**Table 1.** Baseline characteristics of participants with new-onset stroke.

| Characteristics<sup>a</sup> | ELSA | CHARLS |
| --- | --- | --- |
| Sample No. | 99 | 163 |
| Age | 69.3±7.0 | 61.0±8.5 |
| Body mass index, kg/m<sup>2</sup> | 28.2±4.4 | 24.1±4.3 |
| Male | 55 (55.6%) | 79 (48.5) |
| Education | 33 (33.3%) | 14 (8.6) |
| Married | 71 (71.7%) | 146 (89.6) |
| Current smoking | 16 (16.2%) | 75 (46.0) |
| ≥1 alcoholic drink per week | 59 (59.6%) | 42 (25.8) |
| Moderate-vigorous activity | 81 (81.8%) | / |
| Depressive symptoms | 9 (4.2%) | 51 (31.3) |
| Diabetes | 10 (10.1%) | 19 (11.7) |
| Cancer | 7 (7.1%) | 0 (0.0) |
| Chronic lung diseases | 5 (5.1%) | 20 (12.3) |
| Coronary heart disease | 18 (18.2%) | 26 (16.0) |

<sup>a</sup>Education level is defined as ≥NVQ3/GCE A level in ELSA and ≥High school in CHARLS; Heart disease refers to coronary heart disease in ELSA and was not specifically defined in CHARLS.
Abbreviations: ELSA, the English Longitudinal Study of Ageing; CHARLS, the China Health Retirement Longitudinal Study.

In the ELSA cohort, 66 stroke survivors had hypertension before stroke onset, defined as BP≥140/90 mmHg or antihypertensive medication use in at least one survey wave before stroke (Figure 2). Of these, 8 (12.1%) participants were not taking antihypertensive medication in post-stroke surveys and maintained consistently controlled BP after stroke, indicating successful discontinuous antihypertensive medication after stroke. In the CHARLS cohort, 110 stroke survivors had hypertension before stroke. Among them, 15 (13.6%) discontinued antihypertensive medication and maintained controlled BP after stroke. Each BP measurements of stroke survivors who successfully discontinued antihypertensive medication are presented in Table 2 and 3.

**Figure 2.**
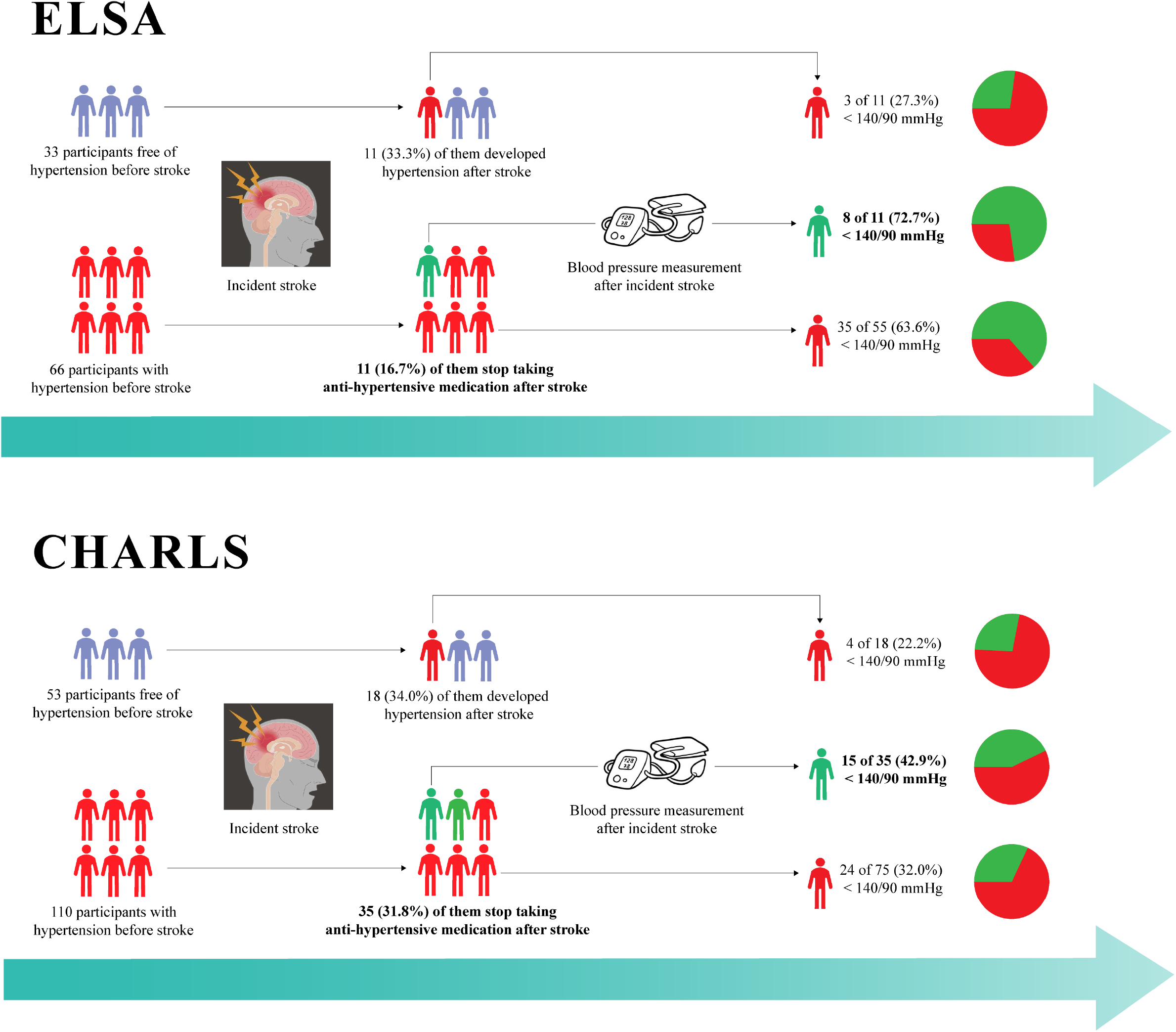
Proportion of stroke survivors with controlled blood pressure after incident stroke.

**Table 2.** Blood pressure measurements of the 8 stroke survivors who successfully discontinued antihypertensive medication after stroke in ELSA.

| Individual ID | wave 4 | Stroke onset | wave 6 | Stroke onset | wave 8 |
| --- | --- | --- | --- | --- | --- |
| 1 | **157/84** |  | **141/84** | ✓ | 138/64 |
| 2 | 138/72 |  | **155/79** | ✓ | 124/87 |
| 3 | **142/56** | ✓ | 126/60 |  | 139/81 |
| 4 | 126/75 | ✓ | 122/69 |  | 126/74 |
| 5 | **155/87** |  | **183/91** | ✓ | 120/61 |
| 6 | 136/87 |  | **148/96** | ✓ | 125/70 |
| 7 | **142/78** | ✓ | 120/77 |  | 127/69 |
| 8 | 115/52 |  | **145/82** | ✓ | 112/72 |

Individual IDs were recoded for privacy. Uncontrolled blood pressure measurements are shown in bold.

**Table 3.** Blood pressure measurements of the 15 stroke survivors who successfully discontinued antihypertensive medication after stroke in CHARLS.

| Individual ID | wave 1 | Stroke onset | wave 2 | Stroke onset | wave 3 |
| --- | --- | --- | --- | --- | --- |
| 1 | **139/92** |  | 129/71 | ✓ | 139/75 |
| 2 | 112/52 |  | **155/86** | ✓ | 123/78 |
| 3 | 120/72 |  | **101/123** | ✓ | 136/82 |
| 4 | **154/88** |  | 115/64 | ✓ | 112/61 |
| 5 | **141/94** |  | **142/96** | ✓ | 134/86 |
| 6 | **141/93** |  | 120/78 | ✓ | 133/85 |
| 7 | **141/76** |  | 134/76 | ✓ | 107/63 |
| 8 | 133/75 |  | 93/57 | ✓ | 111/70 |
| 9 | **155/99** |  | **150/88** | ✓ | 123/78 |
| 10 | 138/72 |  | **151/80** | ✓ | 110/63 |
| 11 | 131/78 |  | 120/73 | ✓ | 127/74 |
| 12 | 110/61 |  | **154/83** | ✓ | 137/68 |
| 13 | 119/80 |  | **145/93** | ✓ | 121/82 |
| 14 | 116/59 |  | 119/62 | ✓ | 131/68 |
| 15 | 128/69 |  | **145/88** | ✓ | 121/88 |

Individual IDs were recoded for privacy. Uncontrolled blood pressure measurements are shown in bold.

The rate of successful antihypertensive medication discontinuation (ceasing medication with maintained BP control) did not differ significantly between the stroke and non-stroke groups, with the crude OR being 0.842 (95% CI: 0.395, 1.800) in ELSA and 1.497 (95% CI: 0.854, 2.623) in CHARLS (Method S2 and Table S2).

Baseline characteristics of participants with pre-stroke hypertension who continued versus discontinued antihypertensive medication after stroke are presented in Table S3. No statistically significant differences were observed in the ELSA cohort. In the CHARLS cohort, participants who discontinued medication had a higher proportion of diabetes (*p*=0.034). At the end of follow-up, among the participants who successfully discontinued medication, two in ELSA had abnormal heart rhythm, and one had angina and myocardial infarction; in CHARLS, one had chronic heart disease (Table S4 and S5).

Among the 55 participants in ELSA who continued antihypertensive medication after stroke, 35 (63.6%) had controlled BP (Figure 2). In ELSA, 11 participants developed new-onset hypertension after stroke. Of these, 3 (27.3%) had controlled BP. The BP control rate among participants with newly diagnosed hypertension post-stroke was significantly lower than that among participants with pre-stroke hypertension (*p*=0.023). In the CHARLS cohort, 22.2% of participants with newly diagnosed hypertension after stroke had controlled BP, but this rate was not significantly different from that of participants with pre-stroke hypertension (32.0%; *p*=0.419).

## Discussion

From three waves of two nationally representative prospective cohorts, ELSA and CHARLS, this study identified 262 participants who experienced a new-onset stroke and had BP measurements before and after the stroke event. In summary, 12.1% to 13.6% of stroke survivors with pre-stroke hypertension successfully maintained controlled BP without antihypertensive medication post-stroke.

To our knowledge, this is the first study to report the proportion of successful discontinuation of antihypertensive medication after a stroke. A key strength of this study is the availability of both pre- and post-stroke BP data, as well as self-reported medication use. The objective BP measurements also facilitated the identification of individuals with unnoted hypertension. The proportion of stroke survivors with controlled BP in the ELSA cohort aligns with findings from two recent studies in the U.S., which reported that approximately 60% to 70% of stroke survivors achieved BP<140/90mmHg.^9,10^

Despite the observation of successful antihypertensive medication discontinuation, the BP management after stroke should not be taken lightly for the following reasons. Firstly, the proportion of stroke survivors who successfully discontinued medication was comparable to that of stroke-free participants. A retrospective study of 2,760 individuals from the general population reported that 42% of hypertensive participant’s BP could return to normal without medication in 6 years.^11^ These findings suggest that medication discontinuation may not be attributed solely to the stroke event.

Secondly, in ELSA, the rate of uncontrolled hypertension was significantly higher among those who had new-onset hypertension after stroke than among those with pre-stroke hypertension. This phenomenon may result from a lack of awareness among individuals without pre-stroke hypertension, leading to an inadequate frequency of BP monitoring after stroke. The group without pre-stroke hypertension may be a key target for improving the post-stroke BP control in England. This difference was not observed in CHARLS, perhaps because the BP control rate in China was lower than that in England.

Finally, this study has certain limitations. One key limitation is that the inclusion of only participants with BP measurements across all three waves may introduce selection bias, as those lost to follow-up were more likely to be older, less educated, and less healthy, limiting generalizability. Additionally, the study did not differentiate between ischemic and hemorrhagic stroke, despite ischemic stroke accounting for approximately 80% of cases.^12^

In summary, 12.1% to 13.6% of stroke survivors with pre-stroke hypertension successfully discontinued antihypertensive medication after stroke. It seems reasonable for clinicians to consider this evidence to support part of “fortunate” stroke survivors to discontinue antihypertensive medication under rigorous BP monitoring. However, BP surveillance remains paramount, especially for stroke survivors with new-onset hypertension after stroke, who are at increased risk of poor BP control after stroke.

## Data Availability

Original survey datasets from the ELSA and the CHARLS are freely available to all researchers. Access to data can be obtained via visiting their websites (https://beta.ukdataservice.ac.uk/datacatalogue/series/series?id=200011, and http://charls.pku.edu.cn/pages/data/111/zh-cn.html). The data can also be obtained on request (Jianian Hua).

## Abbreviations

BP: blood pressure
CHARLS: China Health and Retirement Longitudinal Study
ELSA: English Longitudinal Study of Ageing

## Acknowledgments

The authors thank the U.K. Data Archive and Peking University for providing the data.

## Author Contributions

Jianian Hua and Guochong Chen contributed to the conception and design of the study; Jianian Hua contributed to the acquisition and analysis of data; Qingmei Chen and Jianian Hua contributed to drafting the text or preparing the figures.

## Notes

**Funding Sources** This work was supported by the National Natural Science Foundation of China (Project No. 82202819, Qingmei Chen), Suzhou Science and Technology Planning Project (SKY2022123, Qingmei Chen).

**Conflicts of Interest** None

### Competing Interest Statement

The authors have declared no competing interest.

### Clinical Trial

It is not a clinical trial.

### Funding Statement

This work was supported by the National Natural Science Foundation of China (Project No. 82202819, Qingmei Chen), Suzhou Science and Technology Planning Project (SKY2022123, Qingmei Chen).

### Author Declarations

Ethical approval for ELSA was approved by the London Multicentre Research Ethics Committee (MREC/01/2/91), and CHARLS obtained approval from the Institutional Review Board of Peking University (IRB00001052-11015 for household survey).

